# The adequacy of health system measures in reducing vulnerability to COVID-19 among the health care providers working in primary health care in Rajasthan, India: A Cross-sectional Study

**DOI:** 10.1101/2020.07.21.20149443

**Authors:** Arup Kumar Das, Ambey Kumar Srivastava, Saswata Ghosh, Ruchi Bhargava, Rajan Gupta, Rajesh Ranjan Singh

## Abstract

**Background:** This paper examines the role of individual, facility and system level preparedness in reducing the physiological and psychological vulnerability among primary-level health care providers (HCPs) of COVID19 pandemic in Rajasthan, India.

**Method and Material:** Online and telephonic interviews are conducted among 274 HCPs working in 24 PHCs (17 rural and 7 urban), across 13 districts of Rajasthan. Five dimensions of vulnerability covering awareness, exposure to infection (daily contact; contact with high-risk individuals), physical and mental health conditions, while three aspects of preparedness – at individual (personal care) and facility (provider safety; management and supervision) level – are measured by employing factor analysis. Generalized ordered logit regression model is used to measure the effect of preparedness on COVID19 related vulnerability.

**Result:** Among the 274 HCPs, majority of the staff are from rural PHCs (76 %), less than 35 years (87%), female (57%) and married (57 %). Almost half have high level exposure to COVID19, with mean contact rate is 90. Overall, 26% have comprehensive knowledge on COVID19, and 32% have any mental health issues. Although more than 70% of HCPs have reported more than one individual level preparedness, mental health measures adopted by the HCPs are comparably low. The facility level preparedness for enhancing safety are high such as social distance (79%) and maintaining record of each visitor (75%). However, management related measures adopted by the PHCs are perceived to be lower than the safety measures. The regression analyses suggest that safety related preparedness is significantly associated with reduction of vulnerability by 50%. The management-level preparedness has statistically no significant effect in explaining the variations in level of vulnerability.

**Conclusion:** The facility-level safety measures, which lowers chances of acquiring infection has a positive effect on reducing vulnerability of COVID19. However, the HCPs do not have adequate preparedness at individual, facility management (PHC) and system level to reduce COVID19 vulnerability. Findings suggest that there is a need for a non-conventional approach of monitoring and supervision, in the absence of such measures there is a chance of moral injury that will make the HCPs at the primary level vulnerable to both physiologically and psychologically.

## Introduction

COVID-19 is possibly a novel zoonotic disease (SARS-COV-19), making the entire human population susceptible to infection in the absence of a vaccine [1]. The majority of the transmission happens through contact with droplets of the infected individuals, whether symptomatic or asymptomatic. The disease is considered to be contagious due to the high transmission rate including the asymptomatic phase [2]. There is also evidence of the airborne nature of the disease, particularly among HCPs working closely with COVID-19 infected persons [3]. Health care providers (HCPs) are at the forefront of the fight against the COVID-19 pandemic. Globally, HCPs are considered to be the most vulnerable group, not only in terms of infection and mortality, but also on account of hazards such as prolonged working hours, psychological distress, occupational burnout, stigma, violence [4], and possible transmission to family members [5]. Evidence from China and elsewhere suggests that HCPs face an enormous physiological and psychological vulnerability [6] and stress due to the new norms of wearing personal protective equipment (PPE), leading to hypoxia, hypoglycemia, or sudden cardiac arrest [7, 8, 9, 10]. A study conducted in China suggests that medical health workers are more vulnerable to psychosocial problems than their non-medical counterparts [11]. The study was focused on HCPs working in secondary or tertiary care, engaged directly in managing COVID-19 patients in ICUs and/or ventilation units.

The effects of this pandemic on health care providers working in primary level settings have not been explored in detail. This is of utmost importance in South Asian countries like India and in other lower-income and lower-middle-income countries (LMICs), where primary care is the first level of contact. Nevertheless, HCPs at all levels face common challenges [5], even if of different degrees of severity.

In India, recommendations have been made to engage community health workers, commonly referred as Frontline Health Workers^i^ (FLW), who are the foundation of primary health care [12], in activities related to the prevention of COVID-19. However, the vulnerability and preparedness in the wake of the pandemic faced by the primary health workers including FLWs have not been studied in detail. There is a growing realization that the PHCs have weak preparedness, leading to suboptimal patient safety and infection control measures in the context of COVID-19 [13].

Two important factors make India unique in the context of the current pandemic. First, India has the largest reliance on primary health care (PHCs and Sub-centers), especially in rural areas, which account for about 70% of the Indian population. Second, the pandemic is spreading from urban to rural areas due to the forced return migration after the implementation of the lockdown on 24^th^ March 2020. As a result of this, it is highly likely that rural India would experience outbreaks soon, which will further burden the already overburdened primary health care system with questionable quality of care [14, 15].

The state of Rajasthan, located in the western parts of India, becomes a suitable case to understand the vulnerability and preparedness among primary healthcare providers as a consequence of the COVID-19 pandemic. As the state is 6^th^ in terms of the number of COVID-19 cases (14,691) and the case fatality rate is 23.2 per thousand cases (as on June 21st, 2020) [16], within three months of the start of the pandemic. The state of Rajasthan accounts for 7% of the Indian population, 25% of the population lives in urban areas with a density of 200 population per square KMs and has female literacy of 52% as per the Census 2011. The state is one of the backward states, comes under Empowered Action Group States (EAG) states^ii^, and is lagging behind on several human development indicators compared to the national average (17). The dual burden of communicable and non-communicable diseases is also a major concern for Rajasthan [18, 19]. Although there is no latest available data to understand the situation of the health system in Rajasthan the data available as of 2012-13 highlights major gap in human resource and infrastructure in primary health care facilities [20]. Though Rajasthan has made good progress in improving health outcomes in the recent past, such improvement stands far from achieving the Sustainable Development Goals (SDGs).

The Government of India, the state governments, and various other agencies have come out with many guidelines and have initiated a series of activities to prepare primary health care workers in the wake of the pandemic. These include raising awareness around COVID-19, building capacity, ensuring the availability of PPE, and motivating HCPs to prepare for the eventualities of the COVID-19 outbreak [21]. These policy measures are expected to have a positive effect on the individual, facility, and system-level preparedness so that the vulnerability of HCPs can be reduced.

While individual-level preparedness captures the individual measures adopted by the HCPs to maintain a balanced mental and physical wellbeing, facility-level preparedness captures an individual’s perception about a facility’s readiness in terms of management and safety aspects. Whereas, system-level preparedness is the translation of policy documents into capacity enhancement activities of the HCPs, in the form of training and orientations.

Against this backdrop, the main objectives of this paper are to understand:

1. the vulnerability of the health care workers and the preparedness measures adopted at the individual, facility, and system levels.
2. the effect of the preparedness on reducing vulnerability to the COVID-19 pandemic

## Methods

### Data

A survey among HCPs was carried out between 29^th^ April and 15^th^ May 2020 in 24 PHCs (15 rural and 7 urban) managed by the Lords Education and Health Society (LEHS) through the public-private partnership (PPP) model. These PHCs are spread across 13 of the 33 districts of Rajasthan that account for almost 43% of the total COVID-19 cases, as on June 21, 2020. Functionally, these PHCs are similar to the other government PHCs of Rajasthan, except that they have innovative components such as telemedicine consultants for general as well as specialized care, automatic medicine vending machines, and an additional layer of supervision. Out of the 284 staff working in these facilities, 272 (96.5%) participated in the study. We adopted a combination of the online google forms-based survey and a follow-up telephonic interview to collect the data. We have used a virtual meeting platform to train all interviewers., and randomly selected interviews were monitored for quality assurance. We also obtained ethical approval from the IRB Sigma ethical review board.

### Outcome and explanatory Variables

Box 1 and Box 2 provide a detailed list of variables that were used in computing various dimensions of the outcome variable, the vulnerability; and the primary explanatory variables, which include preparedness at the individual, facility, and system-level respectively. Additionally, some background characteristics of HCPs such as respondent’s age, sex, marital status, living arrangement, and years of experience were included as control variables to measure the adjusted effect of preparedness on vulnerability.

To achieve the study objectives, we explored various dimensions of vulnerabilities (Box 1). First, mixing with the high-risk population, often termed as crowding, is one of the major reasons for transmission [22]. This has been captured by using two variables, namely, the number of individuals the HCPs have contacted in the past 24 hours before the interview, and, among those contacts was there any high-risk population. The next dimension was the HCPs who have comprehensive knowledge of COVID-19. Having comprehensive knowledge helps a person with making better decisions and adopting healthy behaviors. Our definition of a person with comprehensive knowledge was when s/he had correct knowledge on four of the six knowledge items assessed during the survey. Finally, we have also included mental and physical health conditions as measures of vulnerability.

**Box1: Description of the variables used for defining various dimensions of vulnerability**

| Dimensions | Descriptions |
| --- | --- |
| 1 | Individuals who met the 75<sup>th</sup> quintile of the mixing distribution, which was 105 people in the past 24 hours. |
| 2 | staff who had contacted at least three high risk population group (which was 75<sup>th</sup> quintile of the number of high-risk contacts). The High- risk population in this study is defined as those persons or patients who had influenza like symptoms, COVID19 positive person or person who had any direct contact with COVID19 positive cases, people who had any travel history of highly pandemic cities and countries, and persons who are in quarantine. |
| 3 | Individuals who had comprehensive knowledge of COVID19. The respondents were asked to list the persons who are most at the risk of acquiring infection and who are more likely to become critically ill or to die due to the infection. The respondents who were able to tell any four of the following six responses were considered to have comprehensive knowledge.<br>Person who has high risk of acquiring infection: those who travelled to countries or regions which are affected by the pandemic, who has any family member visited countries or regions affected by the pandemic in recent past, migrant or mobile population, health workers who are providing services to COVID19 patients<br>Person who are at risk of being critically ill: Elderly population and persons with any pre-existing chronic illness. |
| 4 | Individuals who had any pre-existing health condition that made them vulnerable to become critically ill due to the COVID19 infection. The pre-exist adverse health conditions included are of pregnancy, diabetic, hypertension and asthma. |
| 5 | Individuals who often experienced any specific mental health issue such as depression, irritation, insomnia, fear, anger, confusion and boredom. |

The preparedness was measured at three levels: (a) at the individual level, it was measured in terms of the personal care measures adopted by a health care provider following the Government of India guidelines. The individual-level measures are the preventive physical and mental health practices adopted by HCPs in line with the Government of India guidelines (https://www.mohfw.gov.in/pdf/PreventiveMeasures.pptx); (b) at the facility level, it was measured in terms of the personal protective or safety measures and management measures put in place for a smooth functioning; (c) at the system level, it was measured through the number of orientations and training sessions conducted by the state health department based on the COVID-19 guidelines laid down by the central as well as the state government. To assess the coverage of the training programs conducted by the health department, we asked each respondent to report the number of COVID-19 training programs they had undergone in the past 3-4 months and the topics covered in those training.

**Box2: Variables included in computing individual and facility level preparedness**

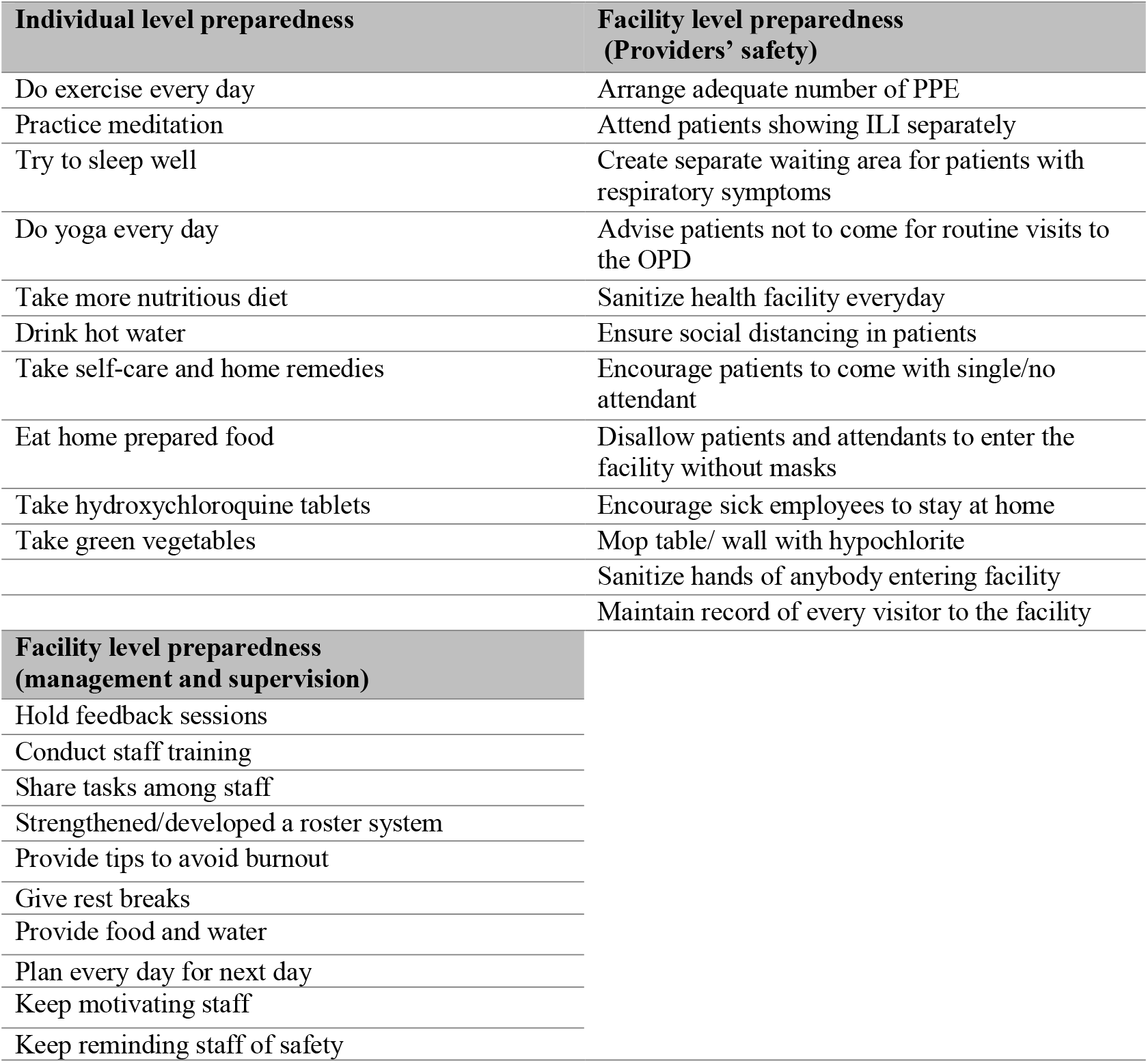

### Statistical analysis

To obtain a summary measure of vulnerability and preparedness, we used factor analysis, with principal factor, to compute the latent measures of vulnerability and preparedness (indices). The factor scores were categorized according to their tercile and classified as high, medium, and low. The first factor was chosen for constructing the indices, which explains maximum variations, with an eigenvalue greater than one. The factor scores of the first factor of vulnerability measures with an eigenvalue of 1.42 were used for computing the vulnerability index as it indicated that the factor had more information than the individual attributes. In the case of individual level preparedness index, the first factor explained 75% of the variation, and all coefficients were found to indicate a positive effect of each of the variables on the factor score. Similarly, the first factor for the two types of facility-level preparedness explained 92% and 98% of the total variation for safety and management indices respectively.

To understand the effect of various preparedness measures on vulnerability to COVID-19, we first used an ordered logit regression model. One of the main problems with such a model is the restriction of the parallel lines assumption, which is often violated. To examine the validity of such an assumption, we have used the brant test with the null hypothesis that the parallel lines assumption holds. The hypothesis was rejected with the brant test Chi2 value of 15.13, p=0.299. However, a detailed examination of the regression coefficients suggested that most of the coefficients varied greatly across regressions. Accordingly, we have used a generalized ordered logit model to allow some of the coefficients to vary, while others remained constant. We have STATA’s user-written command, gologit2 to employ the model [23]. The model is explained with the following expression:

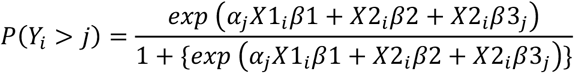

*Where j=1, 2…, M-1*

M is the number of categories in the ordinal variable (vulnerability Index) and *β*’s for X1 and X2 are the same for all values of J but *β*’s for X3 are free to differ. We have used the AIC and BIC criterion for comparing models. We have compared five variations of the model taking into account one of the exposure variables at each time (model 1 to model 4) and finally, a full model (model 5) is created by including all exposure variables, after controlling for background characteristics of HCPs.

## Results

In this section, we elaborate on the variations in vulnerability and preparedness measures across the sub-groups of respondents. Following that, we assess the effect of preparedness on different levels of vulnerability.

### Profile of the respondents

Table 1 provides the socio-demographic and professional details of the 272 staff interviewed in the study. Some of the key observations are that the majority of the staff were young, with 87% of them being less than 35 years old and 57% of them were married. The majority of the staff were working in rural areas (76%) and PHCs (77%). 92% of them were posted in a location that was outside of the district to which they belonged and 48% were living in a rented house. The majority of the staff were either ANM (39%) or Staff Nurse/GNM (20%). More than two-thirds of them (68%) had 2-5 years of work experience, while only 11% had more than 5 years of experience.

**Table 1:** Profile of the respondent staff working in 24 Primary Healthcare Centers (PHCs) Rajasthan, HCP survey, 2020

|  | Percent (%) | Number (N) |
| --- | --- | --- |
| Type of Facility (Rural/Urban) |  |  |
| Rural | 75.7 | 206 |
| Urban | 24.3 | 66 |
| Facility type |  |  |
| Sub-Center (SC) | 22.8 | 62 |
| Primary Health Centre (PHC) | 77.2 | 210 |
| Designation |  |  |
| Medical Officer In-Charge (MOIC) | 8.1 | 22 |
| Pharmacist | 8.1 | 22 |
| Lady Health Visitor (LHV) | 5.9 | 16 |
| Staff Nurse (GNM) | 19.5 | 53 |
| Auxiliary Nurse Midwife (ANM) | 38.6 | 105 |
| Lab Technician (LT) | 7.4 | 20 |
| Multi-Purpose Worker (MPW) | 12.5 | 34 |
| Age category |  |  |
| <=25 yrs. | 23.9 | 65 |
| 25-30 yrs. | 41.5 | 113 |
| 30-35 yrs. | 21 | 57 |
| 35+ | 13.6 | 37 |
| Sex |  |  |
| Male | 42.6 | 116 |
| Female | 57.4 | 156 |
| Marital status |  |  |
| Married | 76.1 | 207 |
| Unmarried | 21.3 | 58 |
| Separated/divorce/other | 2.6 | 7 |
| Residence (Within or outside districts) |  |  |
| Outside district | 91.9 | 250 |
| Within district | 8.1 | 22 |
| Where do you live/ type of residence |  |  |
| Staff quarter | 23.5 | 64 |
| On Rent | 48.2 | 131 |
| Own house | 28.3 | 77 |
| Years of experience |  |  |
| < One yr. | 21.3 | 58 |
| 2-5 yrs. | 67.6 | 184 |
| 5+ yrs. | 11 | 30 |
| Any chronic illness |  |  |
| No | 94.5 | 257 |
| Yes | 5.5 | 15 |
| Source: Authors' calculations from health care providers survey, 2020. |  |  |

### Vulnerability

Table 2 describes variations in the level of vulnerability by selected background characteristics. Overall, 29% of staff belong to the highly vulnerable category with the highest among medical officers (50%) and the lowest among pharmacists (9%). Staff working in sub-centers (34%) were more likely to be highly vulnerable than those working in PHCs (26%). The proportion of staff who belong to the highly vulnerable category are those who belong to the age group of 35+ years (43%), female (30%), separated/divorced (43%), stay in staff quarters (41%). There were also substantial variations across facilities, with the percentage of staff that was highly vulnerable ranging from 9% to 66%.

**Table 2:**
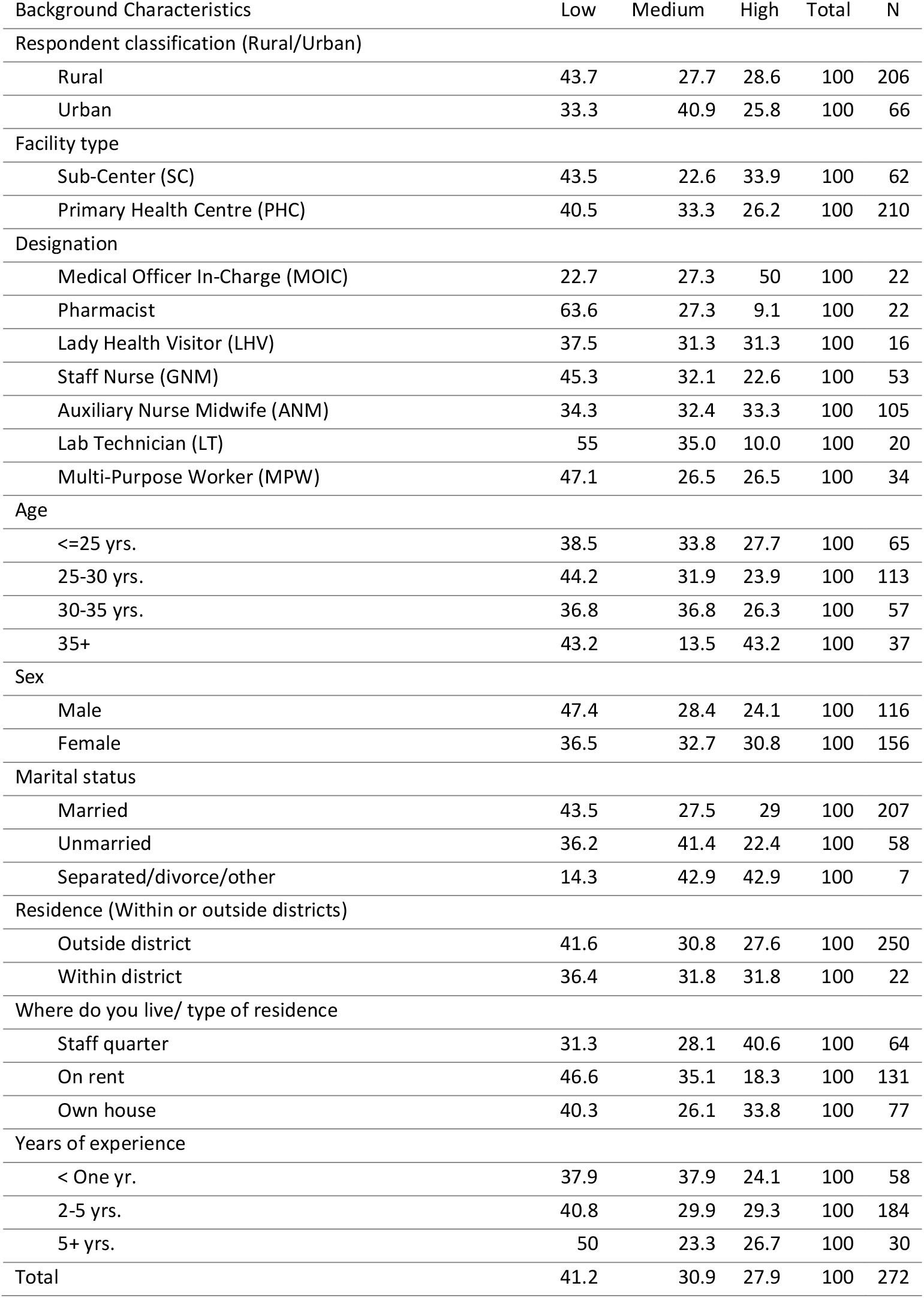
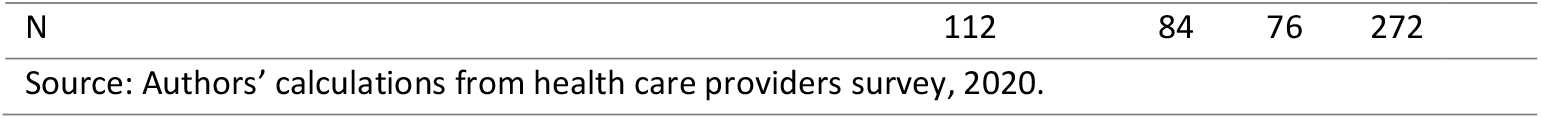
Percent distribution of respondents according to level of vulnerability by background characteristics, HCP survey, 2020

| Background Characteristics | Low | Medium | High | Total | N |
| --- | --- | --- | --- | --- | --- |
| Respondent classification (Rural/Urban) |  |  |  |  |  |
| Rural | 43.7 | 27.7 | 28.6 | 100 | 206 |
| Urban | 33.3 | 40.9 | 25.8 | 100 | 66 |
| Facility type |  |  |  |  |  |
| Sub-Center (SC) | 43.5 | 22.6 | 33.9 | 100 | 62 |
| Primary Health Centre (PHC) | 40.5 | 33.3 | 26.2 | 100 | 210 |
| Designation |  |  |  |  |  |
| Medical Officer In-Charge (MOIC) | 22.7 | 27.3 | 50 | 100 | 22 |
| Pharmacist | 63.6 | 27.3 | 9.1 | 100 | 22 |
| Lady Health Visitor (LHV) | 37.5 | 31.3 | 31.3 | 100 | 16 |
| Staff Nurse (GNM) | 45.3 | 32.1 | 22.6 | 100 | 53 |
| Auxiliary Nurse Midwife (ANM) | 34.3 | 32.4 | 33.3 | 100 | 105 |
| Lab Technician (LT) | 55 | 35.0 | 10.0 | 100 | 20 |
| Multi-Purpose Worker (MPW) | 47.1 | 26.5 | 26.5 | 100 | 34 |
| Age |  |  |  |  |  |
| <=25 yrs. | 38.5 | 33.8 | 27.7 | 100 | 65 |
| 25-30 yrs. | 44.2 | 31.9 | 23.9 | 100 | 113 |
| 30-35 yrs. | 36.8 | 36.8 | 26.3 | 100 | 57 |
| 35+ | 43.2 | 13.5 | 43.2 | 100 | 37 |
| Sex |  |  |  |  |  |
| Male | 47.4 | 28.4 | 24.1 | 100 | 116 |
| Female | 36.5 | 32.7 | 30.8 | 100 | 156 |
| Marital status |  |  |  |  |  |
| Married | 43.5 | 27.5 | 29 | 100 | 207 |
| Unmarried | 36.2 | 41.4 | 22.4 | 100 | 58 |
| Separated/divorce/other | 14.3 | 42.9 | 42.9 | 100 | 7 |
| Residence (Within or outside districts) |  |  |  |  |  |
| Outside district | 41.6 | 30.8 | 27.6 | 100 | 250 |
| Within district | 36.4 | 31.8 | 31.8 | 100 | 22 |
| Where do you live/ type of residence |  |  |  |  |  |
| Staff quarter | 31.3 | 28.1 | 40.6 | 100 | 64 |
| On rent | 46.6 | 35.1 | 18.3 | 100 | 131 |
| Own house | 40.3 | 26.1 | 33.8 | 100 | 77 |
| Years of experience |  |  |  |  |  |
| < One yr. | 37.9 | 37.9 | 24.1 | 100 | 58 |
| 2-5 yrs. | 40.8 | 29.9 | 29.3 | 100 | 184 |
| 5+ yrs. | 50 | 23.3 | 26.7 | 100 | 30 |
| Total | 41.2 | 30.9 | 27.9 | 100 | 272 |

|  |  |  |  |  |
| --- | --- | --- | --- | --- |
| N | 112 | 84 | 76 | 272 |
| Source: Authors' calculations from health care providers survey, 2020. |  |  |  |  |

### Preparedness

Variations at the individual, facility, and system-level preparedness have been presented in Table 3 and the salient findings are as follows.

**Table 3:** Individual and facility level preparedness by background characteristics, HCP survey, 2020

| Background Characteristics | Individual level preparedness |  |  | Facility level preparedness<br>(Management) |  |  | Facility level preparedness<br>(Safety) |  |  | System<br>level | N |
| --- | --- | --- | --- | --- | --- | --- | --- | --- | --- | --- | --- |
|  | Low | Medium | High | Low | Medium | High | Low | Medium | High | % trained |  |
| Respondent classification (Rural/Urban) |  |  |  |  |  |  |  |  |  |  |  |
| Rural | 37.4 | 39.3 | 23.3 | 37.9 | 36.4 | 25.7 | 36.9 | 33.5 | 29.6 | 60.2 | 206 |
| Urban | 22.7 | 54.5 | 22.7 | 19.7 | 24.2 | 56.1 | 22.7 | 43.9 | 33.3 | 74.2 | 66 |
| Facility type |  |  |  |  |  |  |  |  |  |  |  |
| Sub-Center (SC) | 32.3 | 38.7 | 29.0 | 43.5 | 38.7 | 17.7 | 50.0 | 21.0 | 29.0 | 71.0 | 62 |
| Primary Health Centre (PHC) | 34.3 | 44.3 | 21.4 | 30.5 | 31.9 | 37.6 | 28.6 | 40.5 | 31.0 | 61.4 | 210 |
| Designation |  |  |  |  |  |  |  |  |  |  |  |
| Medical Officer In-Charge (MOIC) | 36.4 | 18.2 | 45.5 | 4.5 | 36.4 | 59.1 | 9.1 | 59.1 | 31.8 | 72.7 | 22 |
| Pharmacist | 50.0 | 50.0 | 0.0 | 54.5 | 31.8 | 13.6 | 45.5 | 27.3 | 27.3 | 36.4 | 22 |
| Lady Health Visitor (LHV) | 31.3 | 56.3 | 12.5 | 18.8 | 37.5 | 43.8 | 25.0 | 43.8 | 31.3 | 93.7 | 16 |
| Staff Nurse (GNM) | 34.0 | 37.7 | 28.3 | 30.2 | 35.8 | 34.0 | 20.8 | 47.2 | 32.1 | 60.4 | 53 |
| Auxiliary Nurse Midwife (ANM) | 26.7 | 45.7 | 27.6 | 40.0 | 27.6 | 32.4 | 41.9 | 29.5 | 28.6 | 77.1 | 105 |
| Lab Technician (LT) | 50.0 | 30.0 | 20.0 | 25.0 | 55.0 | 20.0 | 40.0 | 30.0 | 30.0 | 35.0 | 20 |
| Multi-Purpose Worker (MPW) | 35.3 | 55.9 | 8.8 | 35.3 | 32.4 | 32.4 | 35.3 | 29.4 | 35.3 | 41.2 | 34 |
| Age |  |  |  |  |  |  |  |  |  |  |  |
| <=25 yrs. | 36.9 | 43.1 | 20.0 | 40.0 | 26.2 | 33.8 | 32.3 | 27.7 | 40.0 | 63.1 | 65 |
| 25-30 yrs. | 25.7 | 46.0 | 28.3 | 28.3 | 43.4 | 28.3 | 33.6 | 42.5 | 23.9 | 62.8 | 113 |
| 30-35 yrs. | 35.1 | 43.9 | 21.1 | 33.3 | 31.6 | 35.1 | 31.6 | 36.8 | 31.6 | 66.7 | 57 |
| 35+ | 51.4 | 32.4 | 16.2 | 37.8 | 18.9 | 43.2 | 37.8 | 29.7 | 32.4 | 62.2 | 37 |
| Sex |  |  |  |  |  |  |  |  |  |  |  |
| Male | 43.1 | 39.7 | 17.2 | 26.7 | 37.9 | 35.3 | 27.6 | 38.8 | 33.6 | 46.6 | 116 |
| Female | 26.9 | 45.5 | 27.6 | 38.5 | 30.1 | 31.4 | 37.8 | 34.0 | 28.2 | 76.3 | 156 |
| Marital status |  |  |  |  |  |  |  |  |  |  |  |
| Married | 33.3 | 45.4 | 21.3 | 34.3 | 33.8 | 31.9 | 34.8 | 36.2 | 29.0 | 60.9 | 207 |
| Unmarried | 37.9 | 31.0 | 31.0 | 34.5 | 29.3 | 36.2 | 29.3 | 34.5 | 36.2 | 72.4 | 58 |
| Separated/divorce/other | 14.3 | 71.4 | 14.3 | 0.0 | 57.1 | 42.9 | 28.6 | 42.9 | 28.6 | 71.4 | 7 |
| Residence (Within or outside districts) |  |  |  |  |  |  |  |  |  |  |  |
| Outside district | 33.2 | 43.2 | 23.6 | 32.4 | 34.0 | 33.6 | 33.6 | 35.6 | 30.8 | 62.8 | 250 |
| Within district | 40.9 | 40.9 | 18.2 | 45.5 | 27.3 | 27.3 | 31.8 | 40.9 | 27.3 | 72.7 | 22 |
| Where do you live/ type of residence |  |  |  |  |  |  |  |  |  |  |  |
| Staff quarter | 37.5 | 34.4 | 28.1 | 39.1 | 34.4 | 26.6 | 32.8 | 39.1 | 28.1 | 70.3 | 64 |
| On rent | 32.8 | 40.5 | 26.7 | 32.8 | 28.2 | 38.9 | 35.1 | 30.5 | 34.4 | 71.0 | 131 |
| Own house | 32.5 | 54.5 | 13.0 | 29.9 | 41.6 | 28.6 | 31.2 | 42.9 | 26.0 | 45.5 | 77 |
| *Years of experience* |  |  |  |  |  |  |  |  |  |  |  |
| < One yr. | 24.1 | 48.3 | 27.6 | 34.5 | 31.0 | 34.5 | 25.9 | 37.9 | 36.2 | 65.5 | 58 |
| 2-5 yrs. | 32.6 | 44.0 | 23.4 | 33.7 | 32.1 | 34.2 | 34.8 | 33.2 | 32.1 | 64.1 | 184 |
| 5+ yrs. | 60.0 | 26.7 | 13.3 | 30.0 | 46.7 | 23.3 | 40.0 | 50.0 | 10.0 | 56.7 | 30 |
| Total | 33.8 | 43.0 | 23.2 | 33.5 | 33.5 | 33.1 | 33.5 | 36.0 | 30.5 | 63.6 | 272 |
| N | 92 | 117 | 63 | 91 | 91 | 90 | 91 | 98 | 83 | 272 |  |

Source: Authors' calculations from health care providers survey, 2020.

### Individual-level preparedness

It is evident from Table 3, that overall, 23% of the staff belongs to the category of highly prepared at the individual level. Individual-level preparedness was the highest among the medical officers (46%) and the lowest among the Multipurpose Health Workers (MPW) (9%). It is interesting to note that a higher proportion of staff with less than 5 years of experience was highly prepared (28%) than those who had more than 5 years of experience (13%). Likewise, a higher proportion of staff belonging to the sub-centers was highly prepared (29%) than those who were posted in PHCs. Female (28%), unmarried (31%), and staff from outside districts (24%), living in staff quarters (28%), among others, have a high level of individual preparedness than their counterparts.

### Facility level preparedness

Both measures of facility-level preparedness vary across the profile of the respondents. A higher proportion of respondents from the Urban PHCs (56%) than the rural (26%), and PHCs (38%) than SCs (18%) are highly prepared in terms of management-related preparedness. A higher proportion of medical officers (59%) reported a high level of management preparedness than other staff. The safety-related preparedness did not show such substantial variations across the profile of the respondents (Table 3). We have also examined these measures of preparedness across facilities and found that the proportion of respondents who reported a high level of preparedness varies from 8% to 68% for management related preparedness, and from null to 64% for safety-related preparedness (not shown in the table). The correlation between the two measures of facility-level preparedness is not significant, the correlation coefficient being 0.43.

### System-level preparedness

We found that 63% of the staff had received some training or orientation on a COVID-19 topic, with the average being 1.18 training sessions per staff member. There were substantial variations in the proportion of staff that had received any training (Table 3). A higher proportion of urban staff (74%), sub-center staff (71%), Lady Health Visitors (LHV) (94%), and those who had less than one year of work experience (66%) had received at least one training on COVID-19 related issues. It is important to note that a large proportion of pharmacists (64%), lab-technicians (65%), and MPWs (59%) did not receive any training.

### Role of preparedness on reducing vulnerability

The descriptive analysis suggests that there was no significant association between individual preparedness and level of vulnerability. Preparedness of facilities in terms of management and supervision was also not associated with vulnerability. However, a significant association was observed between the preparedness of facilities in terms of safety measures and the level of vulnerability (Table 4). The proportion of staff who were highly vulnerable reduced monotonically from 35% to 21% as the safety-related preparedness increased from low to high (Chi2=9.67, p<0.05). Similarly, as the number of training sessions increased from ‘no training’ to 3+ training sessions, the proportion of staff who were highly vulnerable reduced from 29% to 22%. However, the association was not statistically significant.

**Table 4:** Association between the various level of preparedness and vulnerability, HCP survey, 2020

|  |  |  |  |  |  |
| --- | --- | --- | --- | --- | --- |
| Preparedness | Low | Medium | High | Total | N |
| *Individual preparedness* |  |  |  |  |  |
| Low | 43.5 | 28.3 | 28.3 | 100 | 92 |
| Medium | 42.7 | 33.3 | 23.9 | 100 | 117 |
| High | 34.9 | 33.3 | 31.7 | 100 | 63 |
| *Pearson $\chi^2(4) = 2.2576$ Pr = 0.688* |  |  |  |  |  |
| *Facility preparedness (Safety-Infection prevention)* |  |  |  |  |  |
| Low | 35.2 | 29.7 | 35.2 | 100 | 91 |
| Medium | 36.7 | 37.8 | 25.5 | 100 | 98 |
| High | 53.0 | 26.5 | 20.5 | 100 | 83 |
| *Pearson $\chi^2(4) = 9.6562$ Pr = 0.047* |  |  |  |  |  |
| *Facility preparedness (management)* |  |  |  |  |  |
| Low | 39.6 | 35.2 | 25.3 | 100 | 91 |
| Medium | 45.1 | 30.8 | 24.2 | 100 | 91 |
| High | 38.9 | 28.9 | 32.2 | 100 | 90 |
| *Pearson chi2(4) = 2.3636 Pr = 0.669* |  |  |  |  |  |
| *No. of training attended on COVID-19* |  |  |  |  |  |
| None | 43.4 | 27.3 | 29.3 | 100 | 99 |
| One | 40.2 | 32.9 | 26.8 | 100 | 82 |
| Two | 34.5 | 38.2 | 27.3 | 100 | 55 |
| Three+ | 47.2 | 30.6 | 22.2 | 100 | 36 |
| *Pearson chi2(6) = 2.9362 Pr = 0.817* |  |  |  |  |  |
| Total | 41.2 | 31.6 | 27.2 | 100 | 272 |
| N | 112 | 86 | 74 | 272 |  |
| Source: Authors' calculations from health care providers survey, 2020. |  |  |  |  |  |

The adjusted odds ratios (AORs) of the generalized ordered logit regression – after adjusting for background characteristics including age, sex, marital status, designation, place of work (SC or PHC), facility location (rural or urban), years of experience, living arrangement, and whether they belonged to the same district where the facility was located or to some other district have been shown in Table 5. The results suggest that facility-level safety measures were highly significant in explaining the variations in the level of vulnerability. The comparison of the five models suggests that the M2 is the best fit with the lowest AIC and BIC values, 582.004 and 668.543, respectively. We have also observed from the model that facility-level safety preparedness has a positive and significant effect on reducing the COVID-19 vulnerability of HCPs working in the primary health setting. Those who belong to the medium and high level of preparedness, in terms of safety measures, are less likely to be highly vulnerable (AOR: 0.54; 95%CI: 0.30-0.97) and (AOR:0.36; 95%CI: 0.19-0.66) respectively, than the reference category. The second-best model, full model (M5), also substantiates the same findings. None of the other exposure variables are statistically significant. Among the background characteristics, the staff designation is found to be significant and it is found that medical officers are more likely to be highly vulnerable, followed by ANM and LHV with a predicted probability of being highly vulnerable as 0.45; 95%CI:0.24-0.73, 0.37; 95%CI:0.24-0.51 and 0.28; 95%CI: 0.08-0.48, respectively (Figure 1)

**Table 5:**
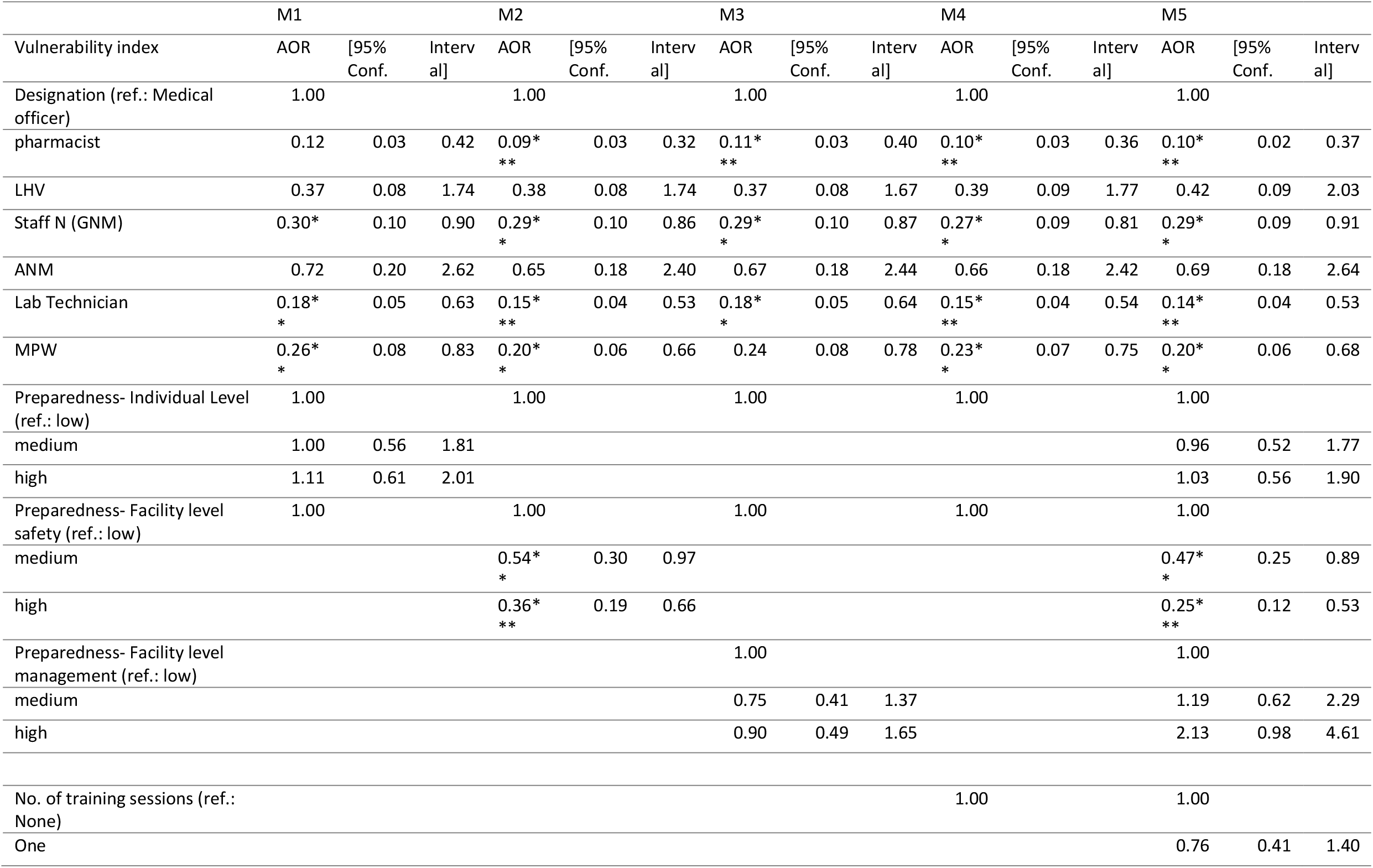
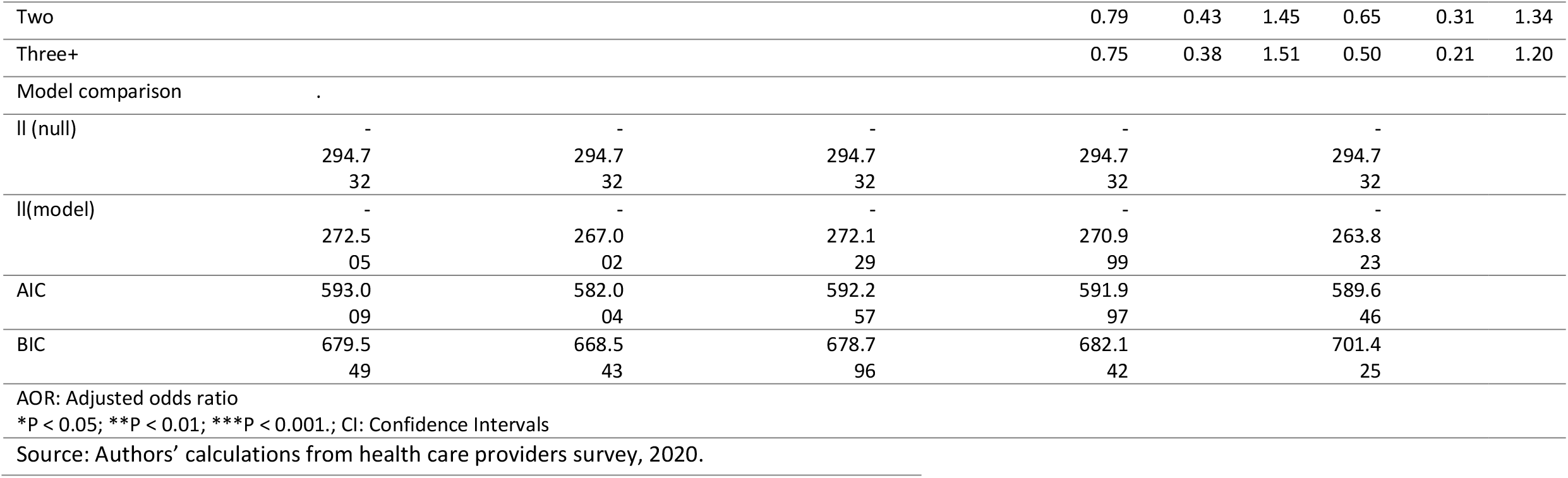
Adjusted odds ratios (AORs) of ordinal logistic regression models and confidence intervals for various level of preparedness on vulnerability index

**Figure 1:**
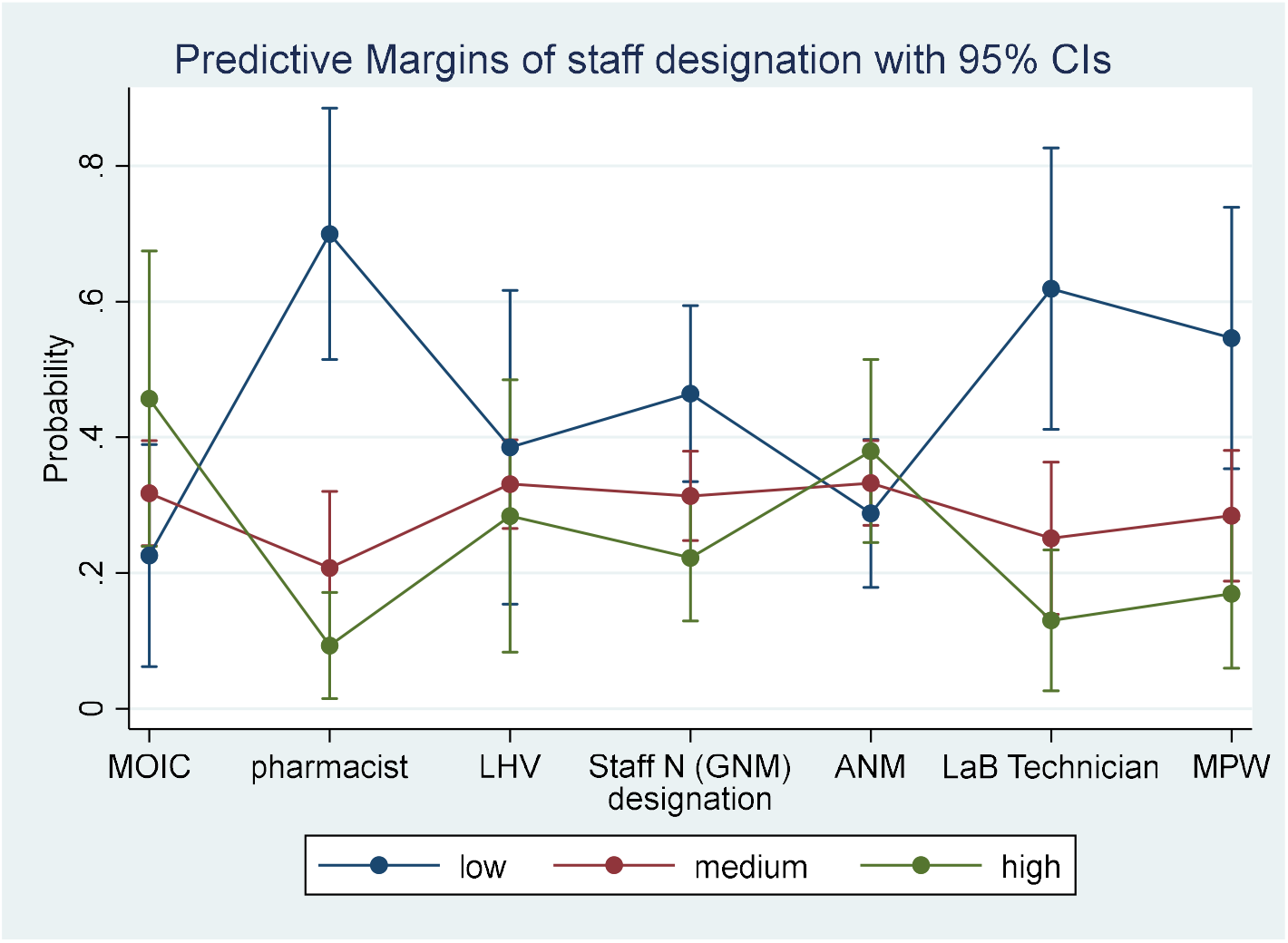
Predicted probability of low, medium and high level of vulnerability by staff designation. **Source: Authors calculation from the health care providers survey**

## Discussion

The present paper highlights the level of vulnerability of HCPs in primary health care vis-à-vis the strength of the support structure to protect their rights. It is emerging that HCPs need to be well-prepared to handle any eventualities which could emerge from the localized outbreak of COVID-19 in the PHC areas by adopting a rights-based approach, as highlighted by the World Health Organisation (WHO) [4, 24]. The facility-level measures directly reduce their chance of acquiring an infection while performing their roles. Inadequate supply of basic equipment in PHCs was a cause for concern across India and in other LMICs even before the COVID-19 pandemic [14].

However, the present study suggests that the HCPs do not have adequate preparedness at the individual level nor the health facility (management mechanisms), and system levels. In the absence of such a support structure, the HCPs are likely to experience more physical and psychological distress. One plausible reason why managerial mechanism at the facility level does not reduce vulnerability could be that there is a need for a non-conventional approach to increasing staff motivation. In the absence of such an approach, there is a possibility of moral injury [25] during the crisis. When a crisis begins, managers should be frank, check on the wellbeing of the staff, provide regular contact, and focus on active monitoring and supervision. By contrast, the data reveals that measures to reduce burnout and to provide personal care to the staff are substantially low. To safeguard the rights, roles, and responsibilities of the HCPs, including sustained social distancing and other safe working environment norms [4], there is a need for redesigning the functioning of the PHCs in terms of infrastructure and human resources and for leveraging innovative technologies like digital health and telemedicine, [26] non-touchable diagnostics, and automated medicine dispenser mechanism, etc. At present, India is facing challenges of efficient supply chain and inadequate stock of PPE [27].

Another important observation is the variations across the 24 facilities is substantial in terms of the vulnerability and preparedness indicators, indicating that there is a need for more localized and micro-planning for a better outcome. Considering the similar health system performance across the EAG states, the findings from the study plausibly can also be used for designing COVID-19 responses in other states.

## Conclusion

As the post COVID-19 era will demand new norms in human society, the service delivery mechanisms will need to be intertwined in the same fashion. In this respect, the study identifies three major areas of change requirements. First, institutionalized systems and mechanisms for highly motivated staff through the alternative method of management and supervision, to prevent them from moral injury. Second, the learning acquired from digital health and telemedicine innovations^iii^ [28, 29] will have a role in maintaining social distance, efficient triaging, referral, and follow-up. Third, it is time to go beyond the dichotomous approach of horizontal and vertical integration and to learn from the experience of the diagonal approach [30] to the health system. All changes have to be customized taking into account concerns of health care providers to prepare the health system to be better equipped for any future crisis.

## Data Availability

Data can be made available on reasonable request to the authors

https://bmcpublichealth.biomedcentral.com/articles/10.1186/s12889-016-3428-8#CR12

## Footnotes

i The front-line health workers incudes the Auxiliary Nurse Midwife (ANM) who are posted at the sub-health center level and responsible for 5000 population, while the Accredited Social Health Activist (ASHA) are village-level volunteers and interphase between community and health system. Usually, there is one ASHA per every 1000 population, as per the norm.

ii There are eight socioeconomically backward states of Bihar, Chhattisgarh, Jharkhand, Madhya Pradesh, Orissa, Rajasthan, Uttaranchal, and Uttar Pradesh, referred to as the Empowered Action Group (EAG) states, lag behind on many of the health and demographic indicators. The maternal mortality ratio is 186 against the India average of 122 and far below from states like Kerala (42) and Tamil Nadu (63). Similarly, the IMR is one of the highest in the country i.e., 38 per thousand live birth, while the same for India is 33 and few other better performing states such as Kerala (10) and Tamil Nadu (16) [Source: Authors compilation from various SRS bulletin]

iii The digital health and telemedicine innovations is one of the successful initiatives in Rajasthan and Delhi implemented WISH [28]. The model is now scaled in Madhya Pradesh in 51 districts, where call centers were initially used for contact tracing, quarantine, and referral management. Similarly, E-*Sanjeevani* (https://esanjeevaniopd.in/About) has become the attraction of general out-patient care services. Such initiatives should be enhanced along with the enhancement of skills and competencies of HCPs in the primary health care sector.

